# Large Language Models Improve the Identification of Emergency Department Visits for Symptomatic Kidney Stones

**DOI:** 10.1101/2024.08.12.24311870

**Authors:** Cosmin A. Bejan, Amy M Reed, Matthew Mikula, Siwei Zhang, Yaomin Xu, Daniel Fabbri, Peter J. Embí, Ryan S. Hsi

## Abstract

**Background:** Recent advancements of large language models (LLMs) like Generative Pre-trained Transformer 4 (GPT-4) have generated significant interest among the scientific community. Yet, the potential of these models to be utilized in clinical settings remains largely unexplored. This study investigated the abilities of multiple LLMs and traditional machine learning models to analyze emergency department (ED) reports and determine if the corresponding visits were caused by symptomatic kidney stones.

**Methods:** Leveraging a dataset of manually annotated ED reports, we developed strategies to enhance the performance of GPT-4, GPT-3.5, and Llama-2 including prompt optimization, zero- and few-shot prompting, fine-tuning, and prompt augmentation. Further, we implemented fairness assessment and bias mitigation methods to investigate the potential disparities by these LLMs with respect to race and gender. A clinical expert manually assessed the explanations generated by GPT-4 for its predictions to determine if they were sound, factually correct, unrelated to the input prompt, or potentially harmful. The evaluation includes a comparison between LLMs, traditional machine learning models (logistic regression, extreme gradient boosting, and light gradient boosting machine), and a baseline system utilizing International Classification of Diseases (ICD) codes for kidney stones.

**Results:** The best results were achieved by GPT-4 (macro-F1=0.833, 95% confidence interval [CI]=0.826–0.841) and GPT-3.5 (macro-F1=0.796, 95% CI=0.796–0.796), both being statistically significantly better than the ICD-based baseline result (macro-F1=0.71). Ablation studies revealed that the initial pre-trained GPT-3.5 model benefits from fine-tuning when using the same parameter configuration. Adding demographic information and prior disease history to the prompts allows LLMs to make more accurate decisions. The evaluation of bias found that GPT-4 exhibited no racial or gender disparities, in contrast to GPT-3.5, which failed to effectively model racial diversity. The analysis of explanations provided by GPT-4 demonstrates advanced capabilities of this model in understanding clinical text and reasoning with medical knowledge.

## 1. INTRODUCTION

Recent progress in generative artificial intelligence (AI) has garnered considerable attention in medical research. AI technologies based on large language models (LLMs) have already demonstrated remarkable capabilities in various clinical applications such as answering questions from simulated medical examinations,^1,2^ complex diagnosis reasoning,^3^ postpartum hemorrhage phenotyping,^4^ and assessing clinical acuity of adults in the emergency department (ED).^5^ However, the potential of integrating generative AI technologies into medical practice remains largely unexplored. There is notable interest from the scientific community not only in further assessing the utility of LLMs for a wide range of clinical applications, but also in studying (1) the mechanisms by which these models reason with medical knowledge, (2) methods to improve LLMs through exposure to medical records during self-supervised training or fine-tuning for specific clinical tasks, (3) strategies to detect and prevent the generation of ‘hallucinations’ that could lead to adverse outcomes in clinical care, and (4) ways to identify and mitigate demographic biases in results generated by LLMs.^6,7^

In this study, we explored the capabilities of three LLMs (*Generative Pre-trained Transformer* 4 [GPT-4], and GPT-3.5, Llama-2)^8,9^ in analyzing ED reports with the specific goal of identifying ED visits due to symptomatic kidney stones. Correct classification of a symptomatic stone event versus one that is not is critically important for understanding disease burden and epidemiology,^10,11^ since acute symptomatic stones are often managed in the ED setting.^12^ In addition, at the patient-level, correct classification of a symptomatic stone event has implications for follow-up care and long-term treatment.^13^ Automatic identification of this phenotype leveraging electronic health record (EHR) data is especially challenging because patients visiting the ED for reasons unrelated to kidney stones often undergo imaging tests such as CT scans and ultrasounds,^10^ which can incidentally reveal kidney stones and may result in a diagnosis of kidney stone disease.^14,15^ Thus, exclusively relying on structured format data for this task, like *International Classification of Diseases*, 9th/10^th^ Revision, Clinical Modification (ICD-9/10-CM) codes during the ED visit−which do not differentiate between symptomatic and asymptomatic kidney stones−may lead to a significant number of false positives. The benefits of solving this task go beyond better phenotyping kidney stone disease for various downstream analyses,^16,17^ as there are many other analogous situations where patients receive secondary diagnoses in critical care settings. Additionally, this research, in conjunction with other related LLM-based phenotyping approaches,^5,18,19^ could be potentially deployed as clinical decision support tools to improve patient triage in the ED.

In our analysis, we implemented several strategies to optimize the performance of LLMs including zero- and few-shot prompting, fine-tuning, and prompt augmentation. We also investigated potential disparities by LLMs with respect to race and gender by implementing fairness assessment and bias mitigation methods for the above-mentioned task. We validated the models using a dataset of ED reports manually reviewed by clinical experts. Based on the explanations provided by the best performing LLM, we further conducted error analysis and assessed its reasoning capabilities in predicting accurate results. Finally, our evaluation includes a comparison between LLMs, traditional machine learning models, and a baseline system utilizing ICD codes for kidney stones.

## 2. METHODS

Figure 1 depicts an overview of the study workflow. First, we describe the process of creating a manually reviewed dataset of ED reports. Next, we explain how we leveraged this dataset to train traditional machine learning models and optimize LLMs for automatically detecting ED reports that indicate kidney stones as the primary reason for the associated ED visit. Finally, during evaluation, we compared these models against a simple baseline system that solves the above-mentioned task using kidney stone-related ICD codes at the time of ED visit. The institutional review board at Vanderbilt University Medical Center (VUMC) approved this study.

**Figure 1.**
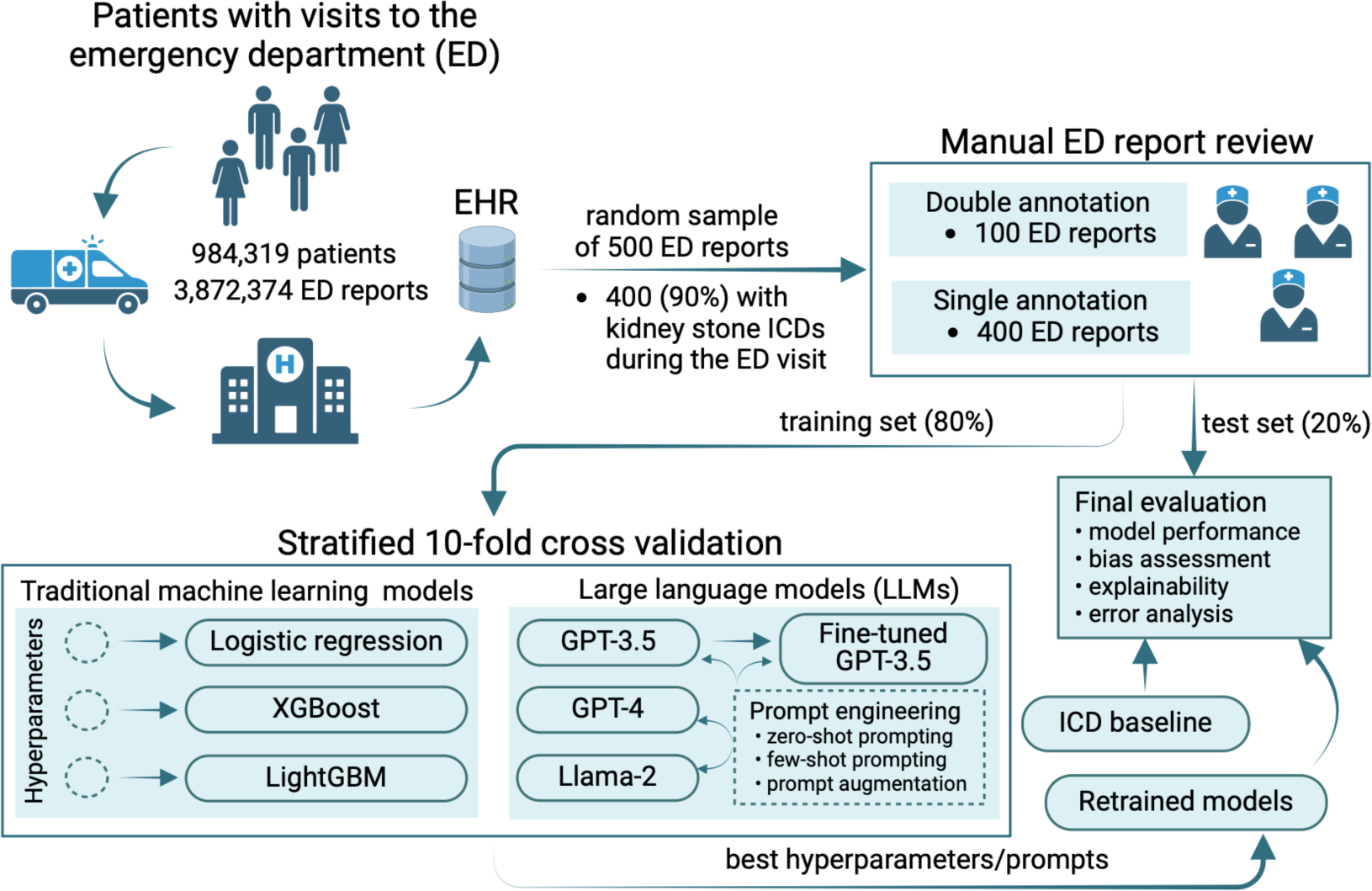
Overview of the study workflow.

### 2.1 Dataset

We retrospectively sampled 500 reports from ∼3.9 million ED reports available in the Synthetic Derivative, a de-identified version of the VUMC’s EHR. Of these, 400 (90%) were randomly sampled from the ED reports associated with a diagnosis of kidney stones as indicated by ICD codes at the time of visit (Figure 1**, Tables S1 and S2**). Three clinical experts (RSH, AMR, and MM) manually reviewed all the selected patient reports and labeled them as positive or negative based on whether they described kidney stones as the primary cause for their ED visit. This process was unbiased as the reviewers were not provided information about which reports were linked to ICD codes for kidney stones. Double annotation was performed on 100 reports and inter-reviewer agreement was measured using Cohen’s kappa statistic. Following the chart review, we divided the reports into training (80%) and test (20%) sets using a stratified random split to maintain the same proportions of positive and negative labels in each set.

### 2.2 Traditional machine learning models

We developed a binary text classification framework using three traditional machine learning algorithms: *logistic regression* (LR), *extreme gradient boosting* (XGBoost), and *light gradient boosting machine* (LightGBM). First, we preprocessed the ED reports using tokenization, lowercasing, and lemmatization, as well as removal of punctuations, numbers, stop words, and tokens of length 1. For the conversion of the preprocessed clinical text into numerical feature vectors, we leveraged the term frequency-inverse document frequency (TF-IDF) weighting scheme. Then, we optimized the hyperparameters for each model using a grid search approach with stratified ten-fold cross validation over the training set. **Table S3** lists all the hyperparameters and their range values selected for optimization. Finally, we retrained the models over the entire training set using the best performing hyperparameter values and conducted the evaluation of these retrained models on the test set. The Python packages used to implement this framework include spaCy (version 3.7.2), scikit-learn (version 1.4.1), xgboost (version 2.0.3) and lightgbm (version 4.3.0).

### 2.3 Large language models

We conducted experiments with three LLMs, namely GPT-4 (version 1106-Preview) and GPT-3.5 (gpt-35-turbo-16k, version 0613) via Azure OpenAI Service, and Llama-2 (Llama 2 70B Chat) through Azure Databricks Model Serving, and assessed their capabilities across four main objectives. The first objective was to evaluate the ability of these models in classifying ED reports (i.e., as either positive or negative for describing kidney stones as the main reason of the ED visit) when provided with prompts of varying specificity in a zero-shot setting. For this, we crafted nine zero-shot prompts that contain a description of the classification task, specific instructions of the output format (e.g., “*The only answer choices are ‘Yes’ or ‘No’*”), additional phenotypic information (e.g., “*the stone is in the kidney or ureter*”), and the ED report to be tested (**Table S4**). Like hyperparameter optimization for the traditional machine learning algorithms, we determined the best performing prompt for each LLM using stratified ten-fold cross validation over the training set.

#### 2.3.1 Ablation studies

The second objective was to conduct ablation studies that could potentially improve the zero-shot prompting results. For the first ablation study, we performed experiments using few-shot prompting based on stratified ten-fold cross-validation on the training set. Particularly, we constructed N-shot prompts (N=1..5) by including randomly sampled ED reports and their manually assigned labels into the zero-shot prompts. In our validation framework, the ED report to be tested was selected from the test fold, while the reports and their labels used as examples in few-shot prompts were sampled from the remaining nine training folds. The second ablation study consisted of augmenting the zero-shot prompts with demographics data and previous history of kidney stone information. This information was extracted from the EHR record associated with the patient at the time of ED visit. **Table S5**, row 1, lists the specific text that was used to augment the zero-shot prompts for this experiment. Finally, for the last ablation study, we tested the potential of achieving better results by exploring LLM fine-tuning capabilities. Specifically, we (1) randomly selected a 9:1 fold split from the same ten-fold cross validation framework over the training set, (2) used the dataset from the nine folds to further train (fine-tune) the GPT-3.5 model for our task, and (3) compared the performance of the fine-tuned model against the initial pre-trained GPT-3.5 model on the reports from the tenth fold. The GPT-3.5 was fine-tuned for 15 epochs using a learning rate multiplier of 0.3. Notably, there were no GPT-4 models available for fine-tuning in our dedicated Azure OpenAI environment.

#### 2.3.2 Bias assessment and mitigation

While recent advancements in generative AI technologies offer promising results for improving clinical care, there is a growing concern that these technologies might introduce and perpetuate biases, possibly resulting in significant harm to various patient categories.^20–24^ To address this concern, our third objective was to investigate the potential racial and gender biases produced by LLMs for our task. The privileged group for studying racial bias consisted of White patients (as opposed to non-White patients), while for studying gender bias, this group was assigned to males (as opposed to females). We conducted bias assessment using two common fairness metrics: *disparate impact* (DI)^25^ and *equal opportunity difference* (EOD).^26^ Briefly, DI measures the ratio of positive label predictions between unprivileged and privileged patient groups. A DI value of 1 indicates no disparity observed in LLM outcomes (with respect to race or gender in our case) while a DI value less than 0.8 suggests potential bias against the unprotected patient group as specified by the ‘*80% rule*’ of disparate impact.^25^ EOD measures the difference in true positive rate (also known as recall or sensitivity) values between unprivileged and privileged patient groups. An EOD value of 0 indicates fairness. To mitigate bias, we used a debiasing technique called *fairness through unawareness*.^27,28^ This approach excludes the explicit use of protected characteristics such as race or gender in the decision-making process.^22,29,30^ In our LLM-based experiments, we applied this technique by omitting race and gender information from prompts, as shown in **Table S5**.

#### 2.3.3 Explainability analysis

The last objective was to gain insights into the reasoning capabilities of LLMs for analyzing ED reports and justifying the classification decisions in solving our task. Specifically, this experiment involved a local explainability analysis, where a clinical expert (RSH) reviewed explanations generated in natural language by GPT-4 for each ED report from the test set. To generate the GPT-4 explanations, we removed the instruction from the prompt that directed the model to give a Yes/No response (e.g., “*Instruction: Choose either ‘Yes’ or ‘No’*”). For the reports where GPT-4 generated correct responses, the review focused on whether its explanations were sound, factually correct, or capable of causing harm. An error analysis was also conducted based on explanations corresponding to false positive and false negative results.

### 2.4. Evaluation setup

We compared the manual annotations against the results extracted by each model and reported performance metrics including precision (positive predictive value), recall (sensitivity), specificity, and F1 score. Because the extraction of positive outcomes was more desirable for our task, we chose F1 as the primary measure in all the optimization experiments during validation (hence, each optimization strategy and ablation study aimed at maximizing the F1 score) and when comparing the models on the test set. Due to the stochastic nature of LLMs,^31^ we repeated all LLM-based experiments ten times and reported the macro-averaged results and their 95% confidence intervals (CIs). To generate predictions with the highest degree of confidence, each LLM was run with the temperature parameter set to 0. We also reported the answer rate for each experiment due to content filters and prompt length limits imposed by LLMs. We evaluated the models with the optimal validated configuration (i.e., the best performing hyperparameter values or prompts) on the test set and compared them to the ICD baseline. To determine whether the differences in F1 values between each model and the ICD baseline are statistically significant, we employed an approximate randomization test based on stratified shuffling^32^ with Bonferroni correction for multiple testing. The python code with all the prompts used in our study is available at https://github.com/bejanlab/LLMs4KS-ED.git.

## 3. RESULTS

### 3.1. Dataset description

From the 500 ED reports (and their corresponding patients), the manual review resulted in identifying 260 (52%) ED visits due to kidney stones and the remaining 240 (48%) visits due to other reasons (**Table 1**). The double-manual review achieved a substantial interrater agreement (Cohen’s κ = 0.78). The patients with ED visits due to kidney stones were younger than the patients with visits due to other reasons (41±17 vs 48±20 years, P=1.58E-05) and had similar gender and race distributions across the two groups. While kidney stone-related ICD codes were assigned to most of the patients with symptomatic kidney stone visits (98.8%), these codes were also assigned to a significant proportion of patients who presented for other reasons (80.4%).

**Table 1.**
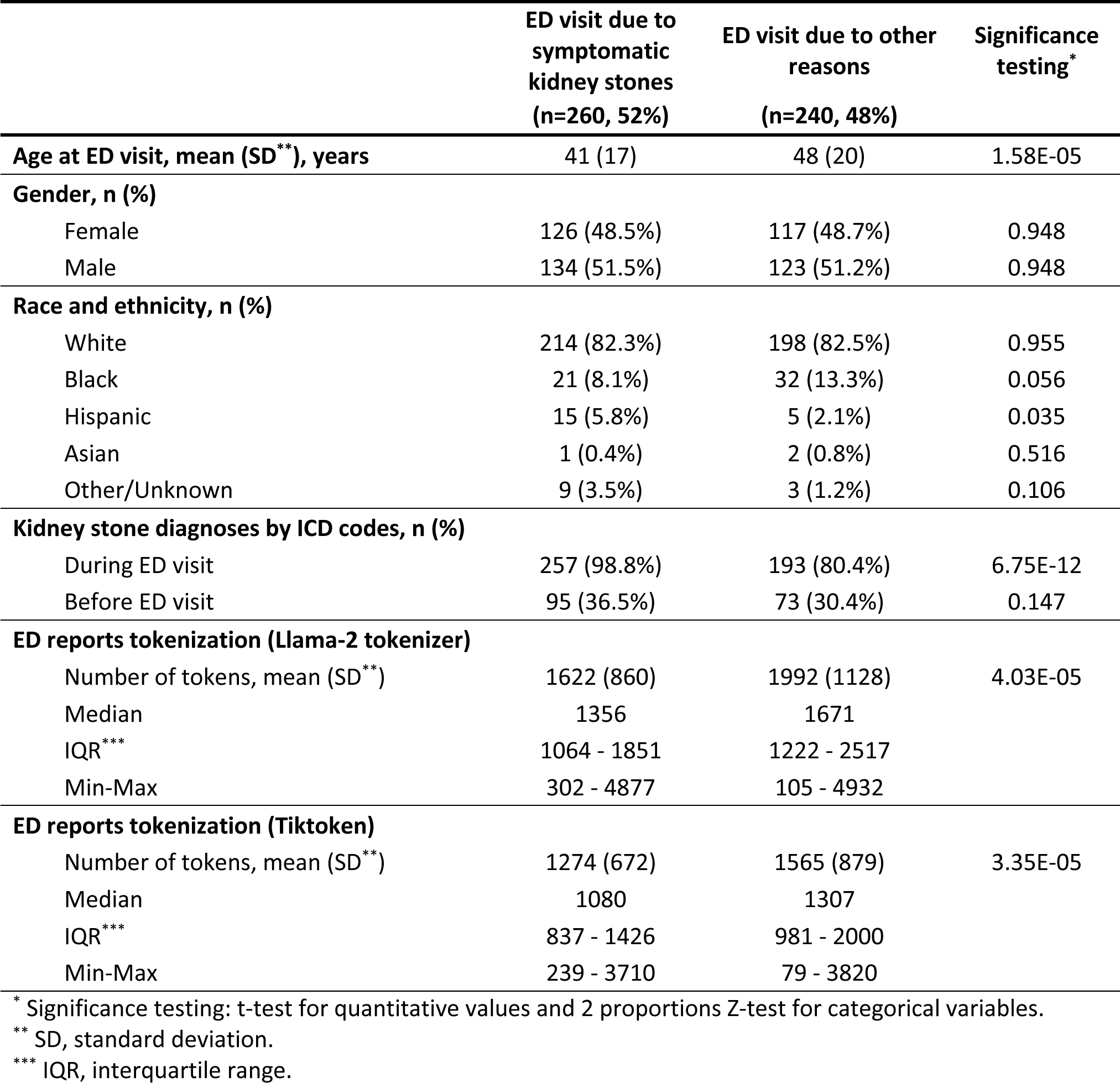
Selected characteristics of study participants and their corresponding ED reports.

|  | ED visit due to<br>symptomatic<br>kidney stones<br>(n=260, 52%) | ED visit due to other<br>reasons<br>(n=240, 48%) | Significance<br>testing* |
| --- | --- | --- | --- |
| **Age at ED visit, mean (SD\*\*), years** | 41 (17) | 48 (20) | 1.58E-05 |
| **Gender, n (%)** |  |  |  |
| Female | 126 (48.5%) | 117 (48.7%) | 0.948 |
| Male | 134 (51.5%) | 123 (51.2%) | 0.948 |
| **Race and ethnicity, n (%)** |  |  |  |
| White | 214 (82.3%) | 198 (82.5%) | 0.955 |
| Black | 21 (8.1%) | 32 (13.3%) | 0.056 |
| Hispanic | 15 (5.8%) | 5 (2.1%) | 0.035 |
| Asian | 1 (0.4%) | 2 (0.8%) | 0.516 |
| Other/Unknown | 9 (3.5%) | 3 (1.2%) | 0.106 |
| **Kidney stone diagnoses by ICD codes, n (%)** |  |  |  |
| During ED visit | 257 (98.8%) | 193 (80.4%) | 6.75E-12 |
| Before ED visit | 95 (36.5%) | 73 (30.4%) | 0.147 |
| **ED reports tokenization (Llama-2 tokenizer)** |  |  |  |
| Number of tokens, mean (SD**) | 1622 (860) | 1992 (1128) | 4.03E-05 |
| Median | 1356 | 1671 |  |
| IQR*** | 1064 - 1851 | 1222 - 2517 |  |
| Min-Max | 302 - 4877 | 105 - 4932 |  |
| **ED reports tokenization (Tiktoken)** |  |  |  |
| Number of tokens, mean (SD**) | 1274 (672) | 1565 (879) | 3.35E-05 |
| Median | 1080 | 1307 |  |
| IQR*** | 837 - 1426 | 981 - 2000 |  |
| Min-Max | 239 - 3710 | 79 - 3820 |  |

\* Significance testing: t-test for quantitative values and 2 proportions Z-test for categorical variables.
\*\* SD, standard deviation.
\*\*\* IQR, interquartile range.

### 3.2. Hyperparameter optimization

Zero-shot prompt optimization identified prompts #9, #7, and #2 as the best performing prompts over the training set for Llama-2 (macro-F1=0.75, 95% CI=0.74−0.76), GPT-3.5 (macro-F1=0.85, 95% CI=0.84−0.87) and GPT-4 (macro-F1=0.86, 95% CI=0.84−0.87), respectively (Figure 2A, **Table S6**). Llama-2 had an 86%–92.5% answer rate due to the model restriction on processing prompts with 4,096 tokens or more, which impacted more ED reports associated with visits due to other reasons (**Table 1**). The answer rate of 99.5% for the GPT models was mainly caused by content filtering policies for self-harm, violence, and sexual content in ED reports (e.g., report mentioning the patient being sexually assaulted). **Table S7** lists the best results obtained by hyperparameter optimization of the traditional machine learning models.

**Figure 2.**
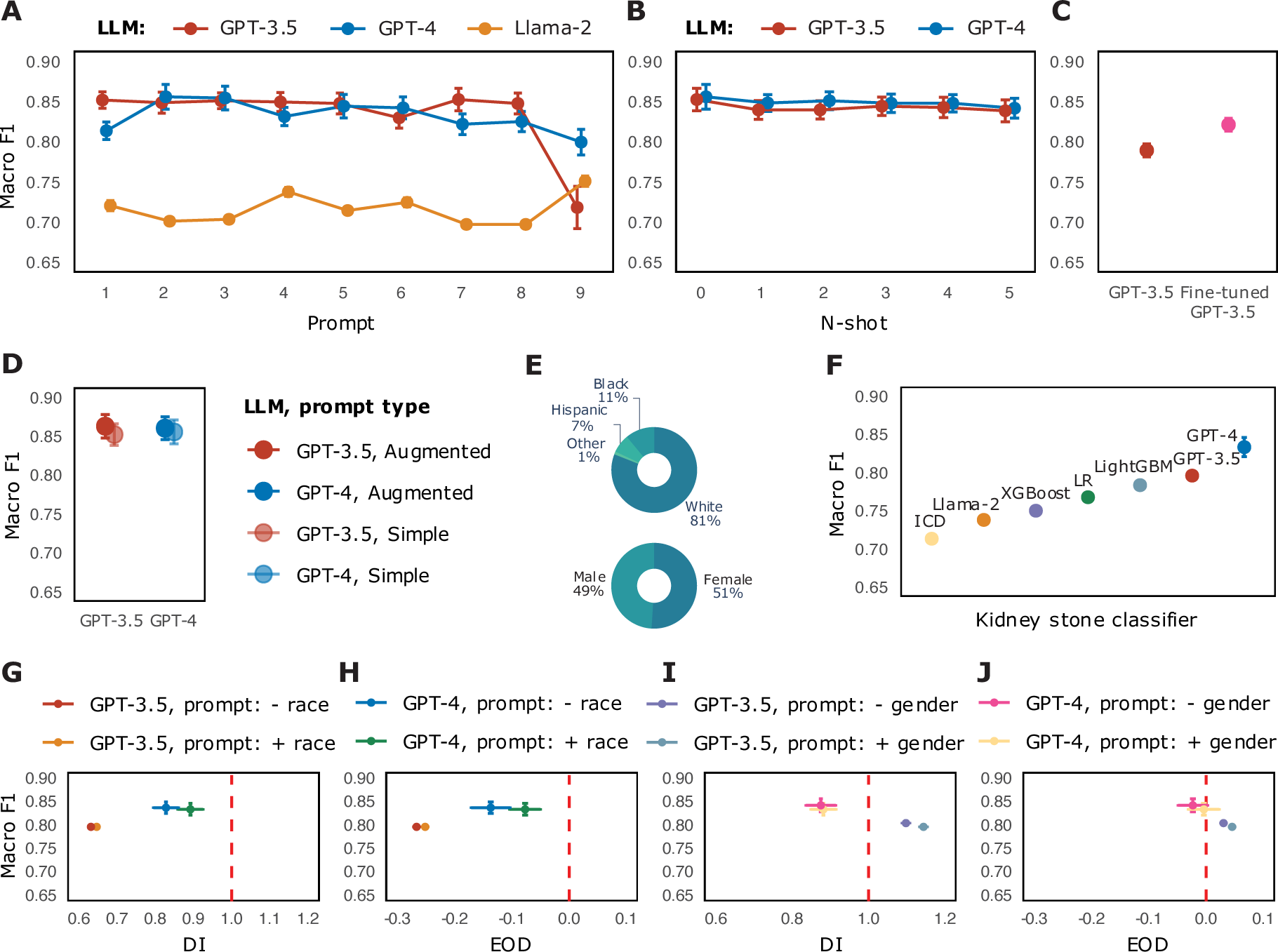
LLM results under various settings. **2A**: Prompt optimization results using zero-shot prompts. Prompts #9, #7, and #2 achieved the best macro-averaged F1 scores for Llama-2, GPT-3.5, and GPT-4, respectively. **2B:** Ablation study results leveraging few-shot prompting (N=1..5). **3C:** Ablation study comparing a fine-tuned GPT-3.5 model against the initial pre-trained GPT-3.5 model. The evaluation was performed on a random fold of a ten-fold cross-validation framework over the training set. **3D:** Ablation study assessing the impact of prompt augmentation with demographic information and previous history of kidney stone. **3E:** Race and gender distribution over the test dataset (N=100). **3F:** Macro-averaged F1 scores achieved by all the models implemented to identify ED visits for symptomatic kidney stones over the test set. **3G-3J:** Racial and gender bias assessment on the test set before and after prompt augmentation with race and gender information. The fairness reference values for disparate impact (DI) and equal opportunity difference (EOD) are 1 and 0, respectively.

### 3.3. Few-shot prompting

Following prompt optimization, we conducted the ablation study based on few-shot prompting by leveraging the highest performing LLMs, GPT-3.5 and GPT-4, and their best zero-shot prompts, #7 and #2, respectively. As shown in Figure 2B and **Table S8**, none of the few-shot experiments outperformed the zero-shot experiments.

### 3.4. LLM fine-tuning

The fine-tuned GPT-3.5 model achieved a macro-F1 of 0.82, substantially higher than the initial pre-trained GPT-3.5 model’s macro-F1 of 0.79 (Figure 2C and **Table S9**). The evaluation was performed on a random fold of the ten-fold cross-validation framework over the training set using the zero-shot prompt #7 (i.e., the best prompt determined by prompt optimization for GPT-3.5). Notably, the GPT-3.5 results for this ablation study are different from the ones obtained during prompt optimization (**Table S6**) because the evaluation was performed under different settings and on different datasets.

### 3.5. Prompt augmentation

The prompt augmentation strategy based on demographic information and kidney stone diagnosis prior to ED visit improved the performance of both GPT-3.5 and GPT-4 models (Figure 3D and **Table S10**). Like the few-shot prompting experiment, this ablation study used the highest performing LLMs and their best zero-shot prompts.

### 3.6. Model evaluation

**Table 2** lists the primary evaluation results for all models on the test dataset, using the optimal parameters derived from hyperparameter tuning, prompt optimization, and ablation studies. Overall, all models achieved superior results when compared to the baseline (Figure 2F). The best results, which were also statistically significantly better than the baseline result, were obtained by GPT-4 (macro-F1=0.833, 95% CI=0.826−0.841, P=7.00E-07), GPT-3.5 (macro-F1=0.796, 95% CI=0.796−0.796, P=1.40E-06), and fine-tuned GPT-3.5 (macro-F1=0.791, 95% CI=0.787−0.795, P=3.50E-06). The P-values, which correspond to approximate randomization tests that compare the difference in F1 scores between each model and the ICD baseline, were adjusted using the Bonferroni correction method. Noteworthy, we evaluated the fine-tuned GPT-3.5 model based on the same prompt configuration we used for fine-tuning (i.e., without prompt augmentation).

**Table 2.** Model evaluation on the test set.

| A Baseline classifier | Optimized hyperparameters |  |  | P | R | S | F1 | AR, % | Significance* |
| --- | --- | --- | --- | --- | --- | --- | --- | --- | --- |
| ICD |  |  |  | 0.5604 | 0.9807 | 0.1667 | 0.7133 | 100 | - |
| B Traditional machine learning algorithms |  |  |  |  |  |  |  |  |  |
| Logistic regression | penalty: l2, C: 1438.45, solver: liblinear |  |  | 0.8085 | 0.7308 | 0.8125 | 0.7677 | 100 | 2.81E-03 |
| XGBoost | n_estimators: 500, max_depth: 4, learning_rate: 0.1, subsample: 0.8, colsample_bytree:1 |  |  | 0.8182 | 0.6923 | 0.8333 | 0.7500 | 100 | 0.1078 |
| LightGBM | n_estimators: 200, max_depth: 5, learning_rate: 0.1, num_leaves: 5 |  |  | 0.8444 | 0.7308 | 0.8542 | 0.7835 | 100 | 6.44E-05 |
| C LLMs | Optimal prompt | N-shot | Prompt augmentation | Macro-P (95% CI) | Macro-R (95% CI) | Macro-S (95% CI) | Macro-F1 (95% CI) | AR, mean (95% CI) | Significance* |
| Llama-2 | 9 | 0 | no | 0.6164<br>(0.616-0.616) | 0.9184<br>(0.918-0.918) | 0.3333<br>(0.333-0.333) | 0.7377<br>(0.738-0.738) | 0.91<br>(0.91-0.91) | 1 |
| GPT-3.5 | 7 | 0 | yes | 0.8478<br>(0.848-0.848) | 0.7500<br>(0.750-0.750) | 0.8542<br>(0.854-0.854) | 0.7959<br>(0.796-0.796) | 1.00<br>(1.00-1.00) | 1.40E-06 |
| GPT-3.5 (fine-tuned) | 7 | 0 | no | 0.8465<br>(0.845-0.848) | 0.7423<br>(0.736-0.748) | 0.8542<br>(0.854-0.854) | 0.7910<br>(0.787-0.795) | 1.00<br>(1.00-1.00) | 3.50E-06 |
| GPT-4 | 2 | 0 | yes | 0.8961<br>(0.888-0.904) | 0.7788<br>(0.769-0.788) | 0.9000<br>(0.892-0.908) | **0.8333**<br>(0.826-0.841) | 0.99<br>(0.99-0.99) | 7.00E-07 |

\* Significance testing: the approximate randomization based on stratified shuffling method was computed to test the difference between the F1 performance values of each classifier and the baseline’s F1 score. *P*-values were adjusted using the Bonferroni correction method. Macro-P, macro-averaged precision (positive predictive value); Macro-R, macro-averaged recall (sensitivity); Macro-S, macro-averaged specificity; Macro-F1, macro-averaged F1-measure; AR, answer rate; CI, confidence interval.

### 3.7 Bias evaluation

We conducted bias assessment and mitigation of the GPT models on the test dataset, which consisted of 100 ED reports corresponding to 81% White and 51% female patients (Figure 2E). The results from **Table 3** and Figures 2G**-2J** show that there were no significant differences in DI and EOD values in either model before and after debiasing (i.e., prompt augmentation with and without race and gender information), nor in model performance. Overall, GPT-4 demonstrated improved DI and EOD values compared to GPT-3.5. However, according to the 80% rule of disparate impact, GPT-3.5 exhibits potential bias against non-White patients (DI=0.64, EOD=-0.26), which remains unresolved despite debiasing efforts.

**Table 3.** Racial and gender bias assessment of GPT-3.5 w/ prompt #7 and GPT-4 w/ prompt #2 before and after prompt augmentation with race and gender information. The fairness reference values for disparate impact and equal opportunity difference are 1 and 0, respectively.

| LLM | Prompt augmentation | DI (95% CI) | EOD (95% CI) | Macro F1 (95% CI) |
| --- | --- | --- | --- | --- |
| **A Racial bias assessment before and after debiasing\*** |  |  |  |  |
| GPT-3.5 | + race | 0.6395 (0.639, 0.639) | -0.2594 (-0.259, -0.259) | 0.7959 (0.796, 0.796) |
|  | - race | 0.6395 (0.639, 0.639) | -0.2594 (-0.259, -0.259) | 0.7959 (0.796, 0.796) |
| GPT-4 | + race | 0.8921 (0.869, 0.915) | -0.0769 (-0.094, -0.060) | 0.8333 (0.826, 0.841) |
|  | - race | 0.8281 (0.805, 0.851) | -0.1370 (-0.161, -0.113) | 0.8369 (0.830, 0.844) |
| **B Gender bias assessment before and after debiasing\*** |  |  |  |  |
| GPT-3.5 | + gender | 1.1438 (1.144, 1.144) | 0.0385 (0.038, 0.038) | 0.7959 (0.796, 0.796) |
|  | - gender | 1.0980 (1.098, 1.098) | 0.0385 (0.038, 0.038) | 0.8041 (0.804, 0.804) |
| GPT-4 | + gender | 0.8818 (0.859, 0.905) | -0.0038 (-0.021, 0.013) | 0.8333 (0.826, 0.841) |
|  | - gender | 0.8751 (0.846, 0.904) | -0.0231 (-0.039, -0.007) | 0.8419 (0.833, 0.851) |

\* Each experiment was repeated 10 times. All the experiments were conducted on the test dataset. DI, disparate impact; EOD, equal opportunity difference; Macro F1, macro-averaged F1-measure; CI, confidence interval.

### 3.8 Assessment of GPT-4 explanations

Most of explanations for GPT-4 predictions were assessed as sound and correct. No explanations were found to be factually incorrect, nonsensical, unrelated to the input prompt, or to cause harm. The explanations conveying uncertainty were properly justified (e.g., “*the definitive diagnosis would depend on the results of the CT scan and urinalysis, which are pending at the time of this report*”). Generally, the disagreements in classification were caused by ED visits for which not enough information was presented in their corresponding reports (e.g., GPT-4 presenting a correct reasoning for a non-kidney stone visit based on the information available in the report, while the clinical expert assessing there is a possibility for kidney stone diagnosis). Two disagreements were identified when the clinical expert assessed kidney stone as the main reason for ED visit while the GPT-4 explanation described a different reason but highly related to kidney stones: (1) dislodged nephrostomy tube, which was previously placed for kidney stones; (2) postoperative complications (constipation and pain) related to a recent kidney stone procedure.

## 4. DISCUSSION

This study explores the abilities of LLMs and traditional machine learning models to analyze ED reports and identify if their corresponding ED visits are due to symptomatic kidney stones. To the best of our knowledge, this is the first study leveraging LLMs for kidney stone phenotyping or for identifying reasons for ED visits. Our evaluation showed that GPT-4 was the best performing model with a macro-F1 of 0.83. The second-best performing results were achieved by GPT-3.5 (macro-F1=0.80). In contrast, Llama-2-70b’s performance (macro-F1=0.74) was inferior to all the models, but remained above the ICD-based baseline (macro-F1=0.71).

The strengths of this study derive from multiple significant findings. First, prompt optimization results confirmed previously reported findings that LLMs are highly sensitive to how prompts are formulated,^23,33,34^ implying that prompt engineering strategies are critical in enhancing LLM-based methods for clinical phenotyping. Even when prompts are semantically similar (e.g., prompt #3 is derived from prompt #2 by substituting ‘emergency department’ with ‘emergency room’), the corresponding performance values achieved by the LLMs vary. Second, no single prompt consistently yields the best results across all models. Our experiments revealed that GPT models performed best with simple and concise prompts (prompts #2 and #7), whereas Llama-2 achieved optimal results with the most specific and complex prompt (prompt #9). Third, despite one of our ablation studies indicated no improvement of few-shot over zero-shot prompting, we believe this study warrants further investigation. One future experiment based on few-shot prompting is to assess all nine prompts instead of only the best zero-shot prompt for each LLM. Another experiment worth considering is applying a similar strategy with retrieval-augmented generation (RAG)^35^ and selecting as examples for few-shot prompts those ED reports that are similar in the embedding space to the ED report being tested. Forth, the fine-tuned GPT-3.5 model outperformed the initial pre-trained GPT-3.5 model when using the same parameter configuration. This finding supports the hypothesis that adapting a pre-trained LLM with clinical data can enhance clinical phenotyping. Fifth, we demonstrated that augmenting the prompts with demographic information and prior disease history helps LLMs make more accurate decisions. We believe this strategy can be applied to a wide range of clinical phenotyping tasks since the patient data used for prompt augmentation can be easily accessed from EHR. Sixth, the bias evaluation revealed that GPT-4 showed no racial or gender disparities whereas GPT-3.5 did not adequately model racial diversity for our task. The overall results suggest that GPT-4 exhibits better fairness measures than GPT-3.5. However, further investigation needs to be conducted on bias mitigation as demographic information may still be encoded in ED reports and, consequently, in their corresponding prompts. Finally, the analysis of explanations provided by GPT-4 indicate advanced capabilities of this model to understand clinical text and reason with medical knowledge. To increase the trustworthiness of using LLMs in clinical settings, we advocate for this type of explainability analysis to become standard practice.

We believe this study achieves progress in understanding the capabilities of LLMs for identifying reasons for ED visits and better phenotyping symptomatic kidney stones. However, efforts towards this goal should persist to ensure the deployment of effective, transparent, and equitable LLM approaches in medicine. For example, the generalizability of our approach should be demonstrated through external validation on larger datasets and with a more heterogeneous study population. Further, additional bias mitigation methods should be implemented to better assess fairness and equity of LLMs for our task.

## Data Availability

Data contain protected health information and are not publicly available. The summary statistics extracted from the EHR data used in this study are provided in the manuscript and supplementary material.

## ACKNOWLEDGMENTS

This research was supported by the National Institutes of Health (NIH) grants R21DK127075, R21HD113234, and UL1TR002243. The authors are also deeply grateful to Chris and Helga Holland for their generous donation, which played a crucial role in facilitating this study. The NIH were not involved in the design and conduct of the study; collection, management, analysis, and interpretation of the data; preparation, review, or approval of the manuscript; or decision to submit the manuscript for publication. The content is solely the responsibility of the authors and does not necessarily represent the official views of the National Institutes of Health.

