## Supplemental Tables for "Large Language Models Improve the Identification of Emergency Department Visits for Symptomatic Kidney Stones"

**Table S1** ICD-9-CM codes for kidney stones.

**Table S2** ICD-10-CM codes from kidney stones.

**Table S3** Hyperparameter values used for optimizing the traditional machine learning models.

**Table S4** Zero-shot prompts for determining if a given ED report indicates kidney stones as the primary reason for the associated ED visit.

**Table S5** Text templates used for prompt augmentation.

**Table S6** Prompt optimization results using zero-shot prompting.

**Table S7** Best results obtained by hyperparameter tuning of the traditional machine learning models.

**Table S8** Ablation study results based on few-shot prompting.

**Table S9** Ablation study results based on the fine-tuned GPT-3.5 model.

**Table S10** Ablation study results based on prompt augmentation with demographic information and previous history of kidney stones.

### Supplemental Tables

**Table S1** ICD-9-CM codes for kidney stones.

| ICD9CM | Description |
|---|---|
| 592 | Calculus of kidney and ureter |
| 592.0 | Calculus of kidney |
| 592.1 | Calculus of ureter |
| 592.9 | Urinary calculus, unspecified |

**Table S2** ICD-10-CM codes from kidney stones.

| ICD10CM | Description |
|---|---|
| N20 | Calculus of kidney and ureter |
| N20.0 | Calculus of kidney |
| N20.1 | Calculus of ureter |
| N20.2 | Calculus of kidney with calculus of ureter |
| N20.9 | Urinary calculus, unspecified |

**Table S3** Hyperparameter values used for optimizing the traditional machine learning models.

| Model hyperparameter | Range |
|---|---|
| **Logistic regression** | |
| penalty | [l1, l2, elasticnet] |
| C | sequence of 20 evenly spaced numbers on the logarithmic scale from [$10^{-4}$, $10^{4}$] |
| solver | [ lbfgs, liblinear ] |
| **XGBoost** | |
| n_estimators | [100, 200, 500] |
| max_depth | [3, 4, 5] |
| learning_rate | [0.01, 0.1, 0.2] |
| subsample | [0.8, 1.0] |
| colsample_bytree | [0.8, 1.0] |
| **LightGBM** | |
| learning_rate | [0.001, 0.01, 0.1] |
| max_depth | [3, 5, 7] |
| n_estimators | [100, 150, 200] |
| num_leaves | [5, 20, 31] |

**Table S4** Zero-shot prompts for determining if a given ED report indicates kidney stones as the primary reason for the associated ED visit.

| No. | Prompt |
|---|---|
| 1 | **System:** You are a clinical expert in identifying symptomatic kidney stone or nephrolithiasis from emergency department reports. Instructions: The only answer choices are 'Yes' or 'No'!<br>**User:** Does the following emergency department report describe a symptomatic kidney stone or nephrolithiasis?<br>Emergency department report: <content> |
| 2 | **System:** As a clinical expert, your role is to determine if the patient's visit to the emergency department is primarily due to experiencing a symptomatic kidney stone or nephrolithiasis. Instruction: Choose either 'Yes' or 'No'.<br>**User:** Is the main cause of the emergency department visit indicated in the following report due to a symptomatic kidney stone or nephrolithiasis?<br>Emergency department report: <content> |
| 3 | **System:** As a clinical expert, your role is to determine if the patient's visit to the emergency room is primarily due to experiencing a symptomatic kidney stone or nephrolithiasis. Instructions: Choose either 'Yes' or 'No'.<br>**User:** Is the main cause of the emergency room visit indicated in the following report due to a symptomatic kidney stone or nephrolithiasis?<br>Emergency room report: <content> |
| 4 | **System:** As a clinical expert, your task is to ascertain whether the patient's emergency department visit is caused by experiencing a symptomatic kidney stone or nephrolithiasis. Instructions: Respond with either 'Yes' or 'No'.<br>**User:** Does the emergency department report specify that the primary reason for the visit is a symptomatic kidney stone or nephrolithiasis?<br>Emergency department report: <content> |
| 5 | **System:** Your expertise lies in determining if the patient's emergency department visit is mainly a result of experiencing a symptomatic kidney stone or nephrolithiasis. Instructions: Select either 'Yes' or 'No'.<br>**User:** In the following emergency department report, is the primary cause of the visit attributed to a symptomatic kidney stone or nephrolithiasis?<br>Emergency department report: <content> |
| 6 | **System:** In your clinical capacity, you are tasked with identifying whether the primary reason for the patient's emergency department visit is the experience of a symptomatic kidney stone or nephrolithiasis. Instructions: Choose 'Yes' or 'No.'<br>**User:** Is a symptomatic kidney stone or nephrolithiasis cited as the main reason for the emergency department visit in the following report?<br>Emergency department report: <content> |
| 7 | **System:** You are a clinician tasked with analyzing emergency department reports.<br>**U:** Given the following emergency department report, output 'Yes' if the reason for the encounter is due to a symptomatic kidney stone or nephrolithiasis. Otherwise, output 'No'. The stone could be in the kidney or ureter.<br>Emergency department report: <content> |
| 8 | **System:** You are a clinician tasked with analyzing emergency department reports.<br>**User:** Given the following emergency department report, output 'Yes' if the reason for the encounter is due to a symptomatic stone in the urinary tract, especially if the stone is in the kidney or ureter. Otherwise, output 'No'.<br>Emergency department report: <content> |
| 9 | **System:** Your responsibility is to assess emergency department reports and ascertain whether the primary reason for the related encounters is due to a symptomatic stone in the urinary tract. A prior definition of this is based on the ROKS nomogram criteria (Rule et al, JASN, 2014). The criteria were: 1) the patient presented for clinical care with gross hematuria or pain, 2) a stone was either seen on imaging in a location consistent with partial, complete, or intermittent obstruction (ureter, uretero-pelvic junction, uretero-vesicular junction, kidney pelvis, or lower kidney pole) or there was documentation that it was voided, and 3) no prior symptomatic episodes from a kidney stone confirmed on imaging or voided. The pain could be typical renal colic or atypical (vague nonlocalized abdominal, pelvic, or back pain). Symptomatic lower pole stones required a clinical pattern of intermittent symptoms from intermittent obstruction at the ureteropelvic junction. In addition to obstructing stones, stone episodes were also considered valid if the patient presented with a symptomatic urinary tract infection from a struvite stone, even if the stone was not obstructing. Incidentally discovered asymptomatic kidney stones on imaging, bladder stones, and “suspected stones” (renal colic but no stone ever seen) were not considered valid episodes.<br>**User:** Given the following emergency department report, determine if the reason for the encounter is due to a symptomatic stone in the urinary tract. The only answer choices are 'Yes' or 'No'!<br>Emergency department report: <content> |

**Table S5** Text templates used for prompt augmentation.

| | Demographic data augmentation | Text template with augmented information |
|---|---|---|
| 1 | + race<br>+ gender | The patient, aged [[AGE]], is [[GENDER]], and of [[RACE/ETHNICITY]] race or ethnicity. [[He/She]] [had/had not] been diagnosed with kidney stones before. |
| 2 | - race<br>+ gender | This is a [[YEAR]]-year-old [[GENDER]] patient. [[He/She]] [[had/had not]] been diagnosed with kidney stones before. |
| 3 | + race<br>- gender | This is a [[YEAR]]-year-old patient of [[RACE/ETHNICITY]] race or ethnicity. The patient [[had/had not]] been diagnosed with kidney stones before. |

**Table S6** Prompt optimization results using zero-shot prompting. This process was conducted based on stratified 10-fold cross validation over the training set and each experiment was repeated 10 times.

| LLM | Prompt | Macro-P (95% CI) | Macro-R (95% CI) | Macro-Spec (95% CI) | Macro-F1 (95% CI) | AR (95% CI) |
|---|---|---|---|---|---|---|
| Llama-2 | 1 | 0.5653 (0.558-0.572) | 0.9944 (0.991-0.998) | 0.1212 (0.103-0.139) | 0.7204 (0.714-0.727) | 0.9225 (0.916-0.929) |
| | 2 | 0.5463 (0.542-0.550) | 0.9794 (0.974-0.984) | 0.0697 (0.058-0.081) | 0.7011 (0.698-0.704) | 0.9100 (0.902-0.918) |
| | 3 | 0.5492 (0.545-0.553) | 0.9794 (0.974-0.984) | 0.0756 (0.064-0.087) | 0.7034 (0.700-0.707) | 0.9125 (0.905-0.920) |
| | 4 | 0.5936 (0.588-0.600) | 0.9747 (0.970-0.980) | 0.2297 (0.218-0.241) | 0.7375 (0.732-0.743) | 0.9200 (0.913-0.927) |
| | 5 | 0.5628 (0.558-0.568) | 0.9800 (0.975-0.985) | 0.1297 (0.116-0.144) | 0.7143 (0.711-0.718) | 0.9150 (0.907-0.923) |
| | 6 | 0.5759 (0.569-0.583) | 0.9797 (0.975-0.985) | 0.1602 (0.143-0.177) | 0.7245 (0.719-0.730) | 0.9175 (0.911-0.924) |
| | 7 | 0.5351 (0.533-0.538) | 1.0000 (1.000-1.000) | 0.0000 (0.000-0.000) | 0.6971 (0.695-0.699) | 0.9250 (0.918-0.932) |
| | 8 | 0.5351 (0.533-0.538) | 1.0000 (1.000-1.000) | 0.0000 (0.000-0.000) | 0.6971 (0.695-0.699) | 0.9250 (0.918-0.932) |
| | 9 | 0.6278 (0.621-0.635) | 0.9363 (0.928-0.945) | 0.3506 (0.327-0.374) | **0.7508** (0.744-0.757) | 0.8675 (0.858-0.877) |
| GPT-3.5 | 1 | 0.9119 (0.897-0.926) | 0.8090 (0.791-0.827) | 0.9061 (0.889-0.923) | 0.8516 (0.841-0.862) | 0.9950 (0.993-0.997) |
| | 2 | 0.8783 (0.864-0.893) | 0.8271 (0.809-0.845) | 0.8678 (0.849-0.886) | 0.8484 (0.835-0.861) | 0.9950 (0.993-0.997) |
| | 3 | 0.8342 (0.821-0.847) | 0.8755 (0.860-0.891) | 0.7999 (0.779-0.820) | 0.8507 (0.841-0.861) | 0.9950 (0.993-0.997) |
| | 4 | 0.8501 (0.836-0.864) | 0.8564 (0.838-0.875) | 0.8268 (0.807-0.846) | 0.8492 (0.838-0.861) | 0.9950 (0.993-0.997) |
| | 5 | 0.8759 (0.859-0.892) | 0.8274 (0.811-0.844) | 0.8639 (0.843-0.885) | 0.8474 (0.834-0.860) | 0.9950 (0.993-0.997) |
| | 6 | 0.9234 (0.910-0.937) | 0.7593 (0.742-0.777) | 0.9268 (0.912-0.942) | 0.8295 (0.817-0.842) | 0.9950 (0.993-0.997) |
| | 7 | 0.9119 (0.900-0.924) | 0.8081 (0.787-0.829) | 0.9110 (0.897-0.925) | **0.8522** (0.838-0.866) | 0.9950 (0.993-0.997) |
| | 8 | 0.8759 (0.859-0.892) | 0.8274 (0.811-0.844) | 0.8639 (0.843-0.885) | 0.8474 (0.834-0.860) | 0.9950 (0.993-0.997) |
| | 9 | 0.9905 (0.985-0.996) | 0.5811 (0.551-0.611) | 0.9911 (0.986-0.997) | 0.7181 (0.692-0.744) | 0.9950 (0.993-0.997) |
| GPT-4 | 1 | 0.7491 (0.736-0.762) | 0.8960 (0.881-0.911) | 0.6644 (0.642-0.687) | 0.8134 (0.803-0.824) | 0.9950 (0.993-0.997) |
| | 2 | 0.8625 (0.847-0.878) | 0.8598 (0.837-0.882) | 0.8422 (0.821-0.863) | **0.8556** (0.840-0.871) | 0.9950 (0.993-0.997) |
| | 3 | 0.8597 (0.844-0.875) | 0.8598 (0.838-0.882) | 0.8382 (0.817-0.860) | 0.8541 (0.839-0.869) | 0.9950 (0.993-0.997) |
| | 4 | 0.8506 (0.836-0.865) | 0.8232 (0.805-0.842) | 0.8311 (0.810-0.852) | 0.8311 (0.820-0.842) | 0.9950 (0.993-0.997) |
| | 5 | 0.8450 (0.829-0.861) | 0.8550 (0.833-0.877) | 0.8184 (0.795-0.842) | 0.8440 (0.829-0.859) | 0.9950 (0.993-0.997) |
| | 6 | 0.8120 (0.795-0.829) | 0.8845 (0.866-0.903) | 0.7641 (0.738-0.790) | 0.8419 (0.828-0.856) | 0.9950 (0.993-0.997) |
| | 7 | 0.7630 (0.748-0.778) | 0.8960 (0.880-0.912) | 0.6866 (0.661-0.712) | 0.8216 (0.809-0.834) | 0.9950 (0.993-0.997) |
| | 8 | 0.7612 (0.746-0.777) | 0.9060 (0.892-0.920) | 0.6789 (0.653-0.705) | 0.8249 (0.812-0.837) | 0.9950 (0.993-0.997) |
| | 9 | 0.9373 (0.922-0.953) | 0.7063 (0.685-0.728) | 0.9408 (0.924-0.957) | 0.7993 (0.783-0.815) | 0.9950 (0.993-0.997) |

Macro-P, macro-averaged precision (positive predictive value); Macro-R, macro-averaged recall (sensitivity); Macro-Spec, macro-averaged specificity; Macro-F1, macro-averaged F1-measure; AR, averaged answer rate; CI, confidence interval.

**Table S7** Best results obtained by hyperparameter tuning of the traditional machine learning models.

| Model | Macro-Precision (95% CI) | Macro-Recall (95% CI) | Macro-Specificity (95% CI) | Macro-F1 (95% CI) |
|---|---|---|---|---|
| Logistic regression | 0.8514 (0.824-0.879) | 0.8374 (0.774-0.901) | 0.8382 (0.798-0.878) | 0.8443 (0.805-0.884) |
| XGBoost | 0.8541 (0.819-0.889) | 0.8707 (0.809-0.932) | 0.8384 (0.796-0.881) | 0.8623 (0.818-0.906) |
| LightGBM | 0.8534 (0.807-0.900) | 0.8417 (0.786-0.897) | 0.8384 (0.779-0.898) | 0.8475 (0.804-0.891) |

CI, confidence interval.

**Table S8** Ablation study results based on few-shot prompting. The GPT-3.5 and GPT-4 experiments were conducted based on augmentations of the zero-shot prompts #7 and #2, respectively.

| LLM | N-shot | Macro-P (95% CI) | Macro-R (95% CI) | Macro-Spec (95% CI) | Macro-F1 (95% CI) | AR (95% CI) |
|---|---|---|---|---|---|---|
| GPT-3.5 | 0 | 0.9119 (0.900-0.924) | 0.8081 (0.787-0.829) | 0.9110 (0.897-0.925) | **0.8522** (0.838-0.866) | 0.9950 (0.993-0.997) |
| | 1 | 0.8442 (0.829-0.859) | 0.8417 (0.826-0.858) | 0.8232 (0.804-0.842) | 0.8394 (0.828-0.851) | 0.9950 (0.993-0.997) |
| | 2 | 0.8475 (0.833-0.862) | 0.8364 (0.822-0.851) | 0.8296 (0.812-0.847) | 0.8394 (0.828-0.851) | 0.9950 (0.993-0.997) |
| | 3 | 0.8579 (0.845-0.871) | 0.8354 (0.820-0.851) | 0.8442 (0.828-0.860) | 0.8439 (0.832-0.855) | 0.9950 (0.993-0.997) |
| | 4 | 0.8672 (0.854-0.881) | 0.8249 (0.807-0.842) | 0.8571 (0.841-0.873) | 0.8424 (0.830-0.855) | 0.9950 (0.993-0.997) |
| | 5 | 0.8686 (0.855-0.882) | 0.8154 (0.797-0.834) | 0.8613 (0.845-0.878) | 0.8382 (0.825-0.852) | 0.9943 (0.992-0.996) |
| GPT-4 | 0 | 0.8625 (0.847-0.878) | 0.8598 (0.837-0.882) | 0.8422 (0.821-0.863) | **0.8556** (0.840-0.871) | 0.9950 (0.993-0.997) |
| | 1 | 0.8460 (0.831-0.861) | 0.8579 (0.843-0.873) | 0.8190 (0.798-0.840) | 0.8480 (0.837-0.858) | 0.9950 (0.993-0.997) |
| | 2 | 0.8539 (0.839-0.869) | 0.8552 (0.840-0.871) | 0.8298 (0.808-0.851) | 0.8507 (0.840-0.862) | 0.9950 (0.993-0.997) |
| | 3 | 0.8523 (0.838-0.867) | 0.8500 (0.834-0.866) | 0.8304 (0.810-0.851) | 0.8476 (0.836-0.859) | 0.9950 (0.993-0.997) |
| | 4 | 0.8456 (0.831-0.860) | 0.8572 (0.842-0.873) | 0.8193 (0.799-0.840) | 0.8476 (0.837-0.858) | 0.9950 (0.993-0.997) |
| | 5 | 0.8462 (0.832-0.861) | 0.8453 (0.827-0.864) | 0.8236 (0.803-0.844) | 0.8414 (0.829-0.854) | 0.9950 (0.993-0.997) |

Macro-P, macro-averaged precision (positive predictive value); Macro-R, macro-averaged recall (sensitivity); Macro-Spec, macro-averaged specificity; Macro-F1, macro-averaged F1-measure; AR, averaged answer rate; CI, confidence interval.

**Table S9** Ablation study results based on the fine-tuned GPT-3.5 model.

| LLM | Macro-P (95% CI) | Macro-R (95% CI) | Macro-Spec (95% CI) | Macro-F1 (95% CI) | AR (95% CI) |
|---|---|---|---|---|---|
| GPT-3.5 | 0.8824 (0.882-0.882) | 0.7143 (0.714-0.714) | 0.8947 (0.895-0.895) | 0.7895 (0.789-0.789) | 1.0 (1.0-1.0) |
| Fine-tuned GPT-3.5 | 0.8889 (0.889-0.889) | 0.7619 (0.762-0.762) | 0.8947 (0.895-0.895) | **0.8205** (0.821-0.821) | 1.0 (1.0-1.0) |

Macro-P, macro-averaged precision (positive predictive value); Macro-R, macro-averaged recall (sensitivity); Macro-Spec, macro-averaged specificity; Macro-F1, macro-averaged F1-measure; AR, averaged answer rate; CI, confidence interval.

**Table S10** Ablation study results based on prompt augmentation with demographic information and previous history of kidney stones.

| LLM | Prompt type | Macro-P (95% CI) | Macro-R (95% CI) | Macro-Spec (95% CI) | Macro-F1 (95% CI) | AR (95% CI) |
|---|---|---|---|---|---|---|
| GPT-3.5 | Simple | 0.9119 (0.900-0.924) | 0.8081 (0.787-0.829) | 0.9110 (0.897-0.925) | 0.8522 (0.838-0.866) | 0.9950 (0.993-0.997) |
| | Augmented | 0.8710 (0.856-0.886) | 0.8612 (0.841-0.882) | 0.8576 (0.840-0.875) | **0.8630** (0.848-0.878) | 0.9950 (0.993-0.997) |
| GPT-4 | Simple | 0.8625 (0.847-0.878) | 0.8598 (0.837-0.882) | 0.8422 (0.821-0.863) | 0.8556 (0.840-0.871) | 0.9950 (0.993-0.997) |
| | Augmented | 0.8689 (0.854-0.884) | 0.8640 (0.841-0.887) | 0.8483 (0.827-0.870) | **0.8605** (0.846-0.875) | 0.9950 (0.993-0.997) |

Macro-P, macro-averaged precision (positive predictive value); Macro-R, macro-averaged recall (sensitivity); Macro-Spec, macro-averaged specificity; Macro-F1, macro-averaged F1-measure; AR, averaged answer rate; CI, confidence interval.
